# Enhancing Emergency Care for Persons Living with Dementia: Innovation and Age-friendly Approaches in Three Emergency Departments

**DOI:** 10.64898/2026.08.19.26360807

**Authors:** Karen A Hauser, Nida F. Degesys, Eric D. Isaacs, Marlena Tang, Jeremy Swartzberg, Vasili Panopulos, Ann Marie Martin, Vincent X Liu, David Schlessinger, Nasrin A Samady, Rohan Malhotra, Colleen Plimier, Azadeh Hadadianpour, Mitchel D. Erickson, Todd James, Stephanie Rogers, Julia Adler-Milstein, Robert Thombley, Sarah Rosenthal, Anna R. Harris, James Hardy, Maria Raven, Malini Singh, Candace Kim, Rachel Perry, Elizabeth Clevenger, Cecilia Carvajal, Donell Babino, Alicia Gray, Melina Shapiro, Tiffany Chan, Heather Allore, Daniella Meeker, Debra Tomasino, Elyssa FL Grogan, Anna Pepper, Madelynn Wellons, Ula Hwang

## Abstract

**Background:** Three San Francisco health system emergency departments have developed Geriatric Emergency Department (GED) models of care programs supporting and providing care for emergency department (ED) patients at risk for or living with dementia. Each system recognized: 1) the high proportion of older adult ED patients and those at risk for dementia, 2) the need to identify cognitive impairment in older adult ED patients, 3) the importance of developing approaches to connect older adult ED patients and their care partners with resources and diagnostic specialty services.

**Methods:** We describe how each hospital adopted and implemented pragmatic GED models of care to support and improve care for ED patients at risk or living with dementia. We also report the proportion of ED encounters made by patients with dementia histories and the number of these reached by GED programs.

**Results:** Three San Francisco hospitals (a tertiary care, critical access, and large integrated health system-community ED) independently implemented GED programs to support and enhance emergency care for patients living with dementia. Each uses screening and assessment tools to identify patients at risk for cognitive impairment. Each captures screening and assessment data to facilitate care and resources for post-discharge care, ensuring coordinated transitions and support for older adults. Programs varied by target patient population age and staff and resource allocation to support program goals. Site-specific pathways differed by location, patient populations, and support from geriatrics, emergency medicine, palliative medicine, neurology, psychiatry, pharmacy, referral processes, and/or pastoral care.

**Conclusions:** Developing GED care interventions that facilitate care for patients at risk of or living with dementia is possible and sustainable when the pathway aligns with health system leadership goals through persistent value demonstration, communication, and promotion. Ultimately, developing and disseminating models of GED care is designed to address geriatric syndromes inclusive of dementia care through continuous quality improvement.

**KEY POINTS:**

- This paper demonstrates the feasibility of implementing Geriatric ED programs with targeted support for people living with or at risk for dementia across three distinct health systems in San Francisco. We describe the reach and implementation processes of each program including commonalities, differences, challenges faced, and opportunities for future improvement.
- Creating unique care pathways for older adults and those with cognitive impairment in the emergency department (ED) is often guided by health system resources.
- Assessing cognition and identifying potential dementia is feasible during an ED visit.
- An interprofessional approach to geriatric ED care is essential to sustainability and success.

## INTRODUCTION

A majority of older adults in the United States (US) visit the emergency department (ED) each year (61 out of every 100 persons 65+ years in age), representing over 20% of total ED visits.^1,2^ As the population ages, the lifetime risk of developing dementia also increases such that the incidence of dementia is projected to nearly double in the next 40 years.^3^ After age 55, men have a 35% and women have a 48% risk of developing dementia.^3^ Unfortunately, persons living with dementia (PLWD) are more than twice as likely to visit the ED compared to other community-dwelling, older adults.^4–6^ Concurrently, a visit to the ED is a sentinel encounter for common geriatric syndromes including cognitive change.^6^ ED visits often precede a dementia diagnosis by 6 to 12 months.^7,8^

The ED is an equalizer for access to healthcare as it is one of the few settings where patients of all backgrounds and conditions are seen and cared for, regardless of demography, socioeconomic factors, or conditions.^8,9^ The ED provides both critical care for those with emergent medical needs and primary care for those that otherwise may have limited access to traditional outpatient care. Many EDs across the country are addressing the special care needs of our aging population by transforming routine care processes and adapting Geriatric Emergency Department (GED) programs.^9^ These programs and their accreditation are aligned with the new Centers for Medicare and Medicaid Services (CMS) metrics for Age-Friendly Hospitals.^10^

GED accreditation standards for neurocognitive screening and care coordination are recognized as care processes,^11^ but implementation strategies for different EDs naturally vary in accordance with local needs and context. In San Francisco, 3 Eds initiated GED implementation protocols specifically for older adult patients and those with potential or documented cognitive impairment. Positive risk screenings for various geriatric syndromes can promote early identification of vulnerable ED patients who may need follow-up care coordination, including referrals to primary care or outpatient evaluation by memory care specialists.

The objective of this paper is to present 3 pragmatic, age-friendly models of GED care designed for PLWD in San Francisco, CA, USA. We describe each program’s implementation of Age-Friendly healthcare practices including common challenges, variations, and innovative solutions. While two of these programs have been previously described,^12,13^ this paper will summarize, compare, and contrast the programs, populations reached, challenges, and novel approaches each have taken in their focus on dementia within broader GED care. We describe the population of older adult patients and those with dementia or cognitive impairment diagnoses with visits to these EDs. We also outline how the GED programs evolved their cognitive screening and management strategies to better identify those at risk and their referral processes.

## METHODS

### Design

Observational descriptive overview comparing 3 San Francisco GED programs and their care for PLWD.

### Settings

San Francisco is the second most densely populated US city with 11 EDs serving over 800,000 diverse residents. The 46.9 square mile geographic area allows ambulatory emergency care patients to select an array of EDs in multiple coverage networks. In 2018, 3 San Francisco EDs—Kaiser Permanente San Francisco (KPSF), University of California San Francisco (UCSF), and Zuckerberg San Francisco General (ZSFG) Hospital—applied for competitive funding support from the Dolby Family Foundation. Each received funding to implement their vision of geriatrics and dementia care in the ED. **Table 1** summarizes the overall characteristics of these 3 EDs. **Table 2** describes key components of interventions that each hospital implemented.

**Table 1:** Health System/Emergency Department characteristics for adult (18 years of age or older) ED population. Parentheticals denote time period reflected by associated data in each cell.

| **Site**<br>**Characteristics for**<br>**adult (18+) ED**<br>**population** | **KPSF** | **UCSF (Parnassus)** | **ZSFG** |
| --- | --- | --- | --- |
| Year initiated GED accreditation process | 2023 | 2020 | 2023 |
| Year accredited | Level 1 (4/2024) | Level 1 (9/2021) | Level 3 (4/2024) |
| Year Dolby program initiated | 2022 | 2020 | 2022 |
| Delivery Model and ED Characteristics | Integrated healthcare delivery system | Academic, quaternary care with some managed care | Public Community Hospital; Level 1 Trauma Center |
| # adult ED visits (18+) | 36,931 (2023) | 40,799 (2024) | 66,601 (August 1, 2024 – July 31, 2025) |
| # 65+ in age at the time of ED visit arrival | 12,186 | 14,868 | 12,644 |

**Table 2:** Emergency Department encounter characteristics for older adults (65 years of age or older).

| **Characteristics of ED encounter by patients 65+ years in age** | **KPSF (2023)** | **UCSF (Parnassus) (April 2021 – December 2024)** | **ZSFG (August 1, 2024 – July 31, 2025)** |
| --- | --- | --- | --- |
| Number 65+ in age at the time of ED visit arrival in stated year or period | 12,186 | 52,750 | 12,644 |
| % 65+ of all adult ED visits | 33% | 36.4% | 17.4% |
| Average (SD) number of ED visits during x year made per unique patient 65+ | 1.5 (1.3) | 1.5 (5.6) | 1.63 (1.3) |
| Insurance |  |  |  |
| % Managed Care | 85.7% | 32.1% | 0 |
| % FFS Medicare | 4.2% | 59.5% | 81.1% |
| % Medi-Cal | 10.1% | 6.7% | 11.9% |
| % Uninsured/Self-Pay only | 0% | 1.2% | 1.5% |
| % Commercial | 0% | 5.2% | 0% |
| % other insurance | 0% | 0.3% | 5.5% |
| Sex |  |  |  |
| % Female | 50.2% | 50.1% | 46.3% |
| % Male | 49.8% | 49.8% | 52.1% |
| % Other | 0 | 0 | 1.1% |
| % missing sex | 0 | 0.05% | 0.43% |
| Race |  |  |  |
| % White | 43.3% | 49.9% | 23.0% |
| % Black or African American | 11.5% | 8.4% | 19.9% |
| % Asian | 31.0% | 30% | 26.3% |
| % American Indian or Alaska Native | 0.1% | 0.2% | 0.3% |
| % Native Hawaiian or other Pacific Islander | 0.2% | 0.50% | 0.7% |
| % other/unknown/missing categories | 13.9% | 10.9%<br>(More than 1 race 4.0%, Other 6.1%, Unknown/Declined 0.8%) | 29.8%<br>(Other 25.1%, Multi-racial 4.3%, Declined to answer 0.3%, Unknown 0.03%) |
| Ethnicity |  |  |  |
| % Hispanic | 10.7% | 8.20% | 24.72% |
| % English speaking | 78.9% | 77.5% | 58.3% |

#### Kaiser Permanente San Francisco (KPSF)

The Dolby award allowed KPSF to develop risk assessments and workflows for older ED patients and those at risk of or living with dementia. KPSF’s GED program began a phased implementation in June 2022, reached full implementation in March 2023, and achieved American College of Emergency Physicians (ACEP) Level 1 accreditation in April 2024. KPSF GED patients are primarily identified using the Geriatric Screening Score (GED-SS). This is a novel, automated risk screening tool embedded within the electronic health record that identifies patients who will most likely have increased health care utilization and/or mortality following an ED visit. The KPSF GED care model utilizes a team approach, with ED physicians, nurses, technicians, case managers, and social workers assisting with GED screening and assessments. Two new roles were introduced: a dedicated GED pharmacist (available weekdays for 40 hours per week) for medication review and a Geriatric Clinical Nurse Specialist (GeriCNS, available weekdays on average 32 hours per week) to perform cognitive screening and geriatric assessment. The case management consultation, nursing, and technician workflows for mobility screening, delirium screening, and age-friendly care interventions operate 24/7. As ninety-six percent (96%) of patients in the GED are KPSF members, preventing avoidable hospitalization and managing risks for these patients contributes to the sustainability of the program.

#### University of California San Francisco (UCSF)

The UCSF Helen Diller Medical Center at Parnassus Heights is a 500-bed academic medical center and quaternary care hospital. The UCSF ED initiated a GED program in 2020 and received initial ACEP Level 1 accreditation in 2021. It was developed in cooperation with the Division of Geriatric Medicine and the Department of Neurology. The program utilizes Division of Geriatrics embedded nurse practitioners (NP) with advanced geriatrics training, social workers, and others. The payer-mix for ED patients is approximately one-third commercial, one-third Medi-Cal, and one-third Medicare. The primary goals of the GED program are to reduce hospital admission and associated risks, length of stay, if appropriate, facilitate safe ED discharge, coordinate care transitions to community services, facilitate referrals to the outpatient UCSF Memory and Aging Center (MAC), and improve ED patient experience. The Dolby award allowed the ED to develop and establish its program with a focus on patients with, at risk of, or living with dementia.

#### Zuckerberg San Francisco General (ZSFG)

The ZSFG ED is a county public hospital and level 1 trauma center. ZSFG serves as a safety net medical center, providing the acute medical care, hospital services, and specialty and primary care as part of a delivery network of public health providers and federally qualified health centers in San Francisco. The Dolby award catalyzed the ZSFG GED program that began implementation in 2023 and received ACEP Level 3 accreditation in 2024. Most recently, ZSFG achieved Level 2 GED status in 2025. The award enabled the ED to address health IT, reporting, and analysis needs, add patient navigators and dedicated social workers for older ED patients, and provide screening and linkage to resources for patients at risk of or living with dementia.

See **Table 1** for details about each ED, their respective health systems, and their GED program implementation during the most recent years available.

### Dementia patient population seen in EDs

To estimate the potential reach of their programs among patients with prevalent dementia or cognitive impairment, we characterize two populations: Population 1 includes all ED patients over 65 years of age. Population 2 includes ED patients over 65 years of age with medical diagnoses of dementia or cognitive impairment. We constructed our definition of dementia or cognitive impairment diagnosis codes to be as inclusive as possible by defining an encounter-level indicator of dementia as any qualifying ED encounter where the patient had one or more ICD-10 diagnosis codes for dementia or cognitive impairment^14–16^ in their medical history, problem list, referrals, or billing record available for extraction from health system databases. The complete list of qualifying ICD-10 codes is listed in the appendix.

See **Table 2** for characteristics of geriatric patients in the ED and **Table 3** for ED patients with dementia or cognitive impairment diagnoses.

**Table 3:** ED visits with any ICD-10 dementia or cognitive impairment diagnoses documented a same day or before an ED encounter.

| **Attribute for adult ED population** | **KPSF** | **UCSF**<br>**(Parnassus)** | **ZSFG** |
| --- | --- | --- | --- |
| Encounter period | 2023 | April 2021 –<br>December 2024 | August 1, 2024<br>–<br>July 31, 2025 |
| Length of history for dementia diagnosis | 2001 –<br>index ED<br>encounter | 2001 - index ED<br>encounter | August 2019 -<br>index ED<br>encounter |
| # of all 18+ ED visit encounters with ANY ICD-10 dementia or CI diagnoses prior to and including index encounter | 3,490 | 14,658 | 2,487 |
| # of all 65+ ED visit encounters with ANY ICD-10 dementia or CI diagnoses up to and including index encounter | 3,099 | 12,218 | 1,858 |
| % 65+ ED encounters with previous ICD dementia or CI diagnoses | 25.4%<br>(3099/12,186) | 23.2%<br>(12,218/52,750) | 14.7%<br>(1,858/12,644) |

### GED programs, development of components, and evolution

Each ED developed and implemented their GED programs over different years, with the earliest program starting at UCSF in 2020, to the most recent beginning implementation in 2023 (both KPSF and ZSFG).

### Development of eligibility screening tools and procedures

All three sites began their GED and dementia programs using variants of risk screening tools for eligibility determination for higher-intensity assessment of geriatric or dementia care interventions. There was variation in how processes evolved over time based on health system resources and patient populations. For example, KPSF has the unique ability to aggregate and analyze patients’ historical records, enabling the creation of a flag that can automatically be populated at triage.^13^ In addition to this automated flag, KPSF also includes a manual flag for patients who their ED clinicians subjectively think should be eligible for GED care. Similarly, after relying on the Identification of Seniors at Risk (ISAR) tool,^17^ UCSF has shifted to a “gestalt” frailty assessment question administered by ED triage nurses. All three sites had different approaches with regard to who on the ED care team could direct a patient to GED care. This type of qualitative referral is a recognition of the value of GED protocols, as well as the limitations of standardized instruments in identifying patients who would benefit most from GED care.

#### Workflows and care coordination during encounter

Common approaches to identifying patients to the broader ED care team were used, including wristbands and posted signs with details of patient preferences and dementia care needs. The embedded geriatric NP at UCSF plays a central role in conducting a modified comprehensive geriatric assessment (mCGA) and directing follow up care. The UCSF mCGA is a systematic screening of medical, cognitive, and functional capacities which is utilized to create an integrated, patient centered treatment plan that reflects topics important for older adults.^18^ Meanwhile, KPSF relies on the GENIE role (Geriatric Emergency Nurse Initiative Experts) and ZSFG places substantial responsibility on a patient care navigator.

The ZSFG Age-Friendly ED (AFED) Patient Navigator and Social Worker coordinate with home health agencies; connect patients to meal delivery programs, community meal and nutrition resources, food banks, and frozen or hot meal pick-up locations; assist patients in finding a primary care provider (PCP) and ensuring discharge follow-up appointments are scheduled; order appropriate durable medical equipment (DME); arrange home safety evaluations (e.g. The Community and Home Injury Prevention Program for Seniors); and provide referrals to Adult Day Health Centers and community-based organizations such as senior centers. In addition, the ZSFG AFED team collaborates with an existing Social Medicine service to provide comprehensive care for patients with housing instability, substance use disorders, and the need for comprehensive case management.

At UCSF, the AFED team connects patients to enhanced care management programs, fall prevention exercise classes, Alzheimer’s Association and dementia care services, the Guiding an Improved Dementia Experience (GUIDE) Program through UCSF Care at Home,^19^ and community agency programs. Additional referrals may include resources through the Department of Aging and Disability Services, family caregiver support organizations (e.g., Family Caregiver Alliance, Institute on Aging), private duty nursing agencies, homeless support services (e.g., shelters, showers, laundry facilities, and inclement weather centers), and mental health resources. To address behavioral expression challenges for dementia or delirium, which may interfere with usual care, a care pathway called Code DICE (Describe, Investigate, Clarify, Evaluate) was developed.^20^ This pathway proactively revises the care plan to promote behavioral de-escalation and strengthen patient and staff safety.

### Challenges

#### Kaiser Permanente San Francisco (KPSF)

The KPSF GED team has identified gaps in post ED care regarding confirmatory cognitive testing with Montreal Cognitive Assessment (MoCA) after ED discharge. GED leadership has partnered with outpatient care providers, including community paramedicine, neurology, dementia core committee, and supportive care services to continue improving upon follow-up care for vulnerable older adults with abnormal cognitive screens. Continuing to streamline and define the cognitive screening process in the ED remains a significant challenge with the resource loss of the ED Geriatric CNS (after 2024), and is currently a work in process. Like the other ED programs, KPSF GED is unable to directly refer to the memory clinic routinely but can be done in special cases.

#### University of California San Francisco (UCSF)

The UCSF AFED team has experienced programmatic challenges related to dementia care and resource connections despite the close coordination with the UCSF MAC. The MAC stopped accepting direct referrals from the AFED Team. Instead, referrals were preferred directly from the PCP for a post-ED follow-up appointment secondary to a potential for intervening medical decompensation. The PCP is notified of the mCGA consultation note with the referral details.

Other challenges include the resource loss of the MAC patient navigation assistance and the obstacle of prior authorization with Medicare Advantage Plans. These barriers may limit timely and efficient access to formal cognitive evaluation. Despite these barriers, the AFED team continues to place direct referrals to the MAC and provides navigation assistance when possible. They maintain consult note communication with both neurology (MAC) partners and the patient’s PCP.

Sustained involvement by social workers and care coordinators is limited by workload challenges, which limits care continuity. However, collaboration with memory clinics shows promise for improving access^21^ to formal neuropsychiatric testing and potential therapy options.

#### Zuckerberg San Francisco General (ZSFG)

As a public hospital, ZSFG has limited capacity for new patients in the Geriatrics and Memory Clinics located on the hospital campus. As such, there is some hesitancy to add direct referral from the ED to these specialty clinics unless specific criteria are met. The Geriatric Clinic will accept direct referral of ED patients if their primary care home is in the public health network and they cannot see their PCP within 2 weeks after the ED visit. The ZSFG Neurology Memory Clinic has asked for patients with cognitive impairment to be referred through their PCP.

**Table 4** provides details of the three GED programs, eligibility criteria, how and when patients were identified during the ED visit encounter, which teams evaluated eligible patients, and details of the care processes and referrals received. Of note, two of the GED programs (UCSF and ZSFG) focused on patients 65+ years of age, while the KPSF program focused on patients 70+ years of age.

**Table 4:** GED Intervention Design and Care. Abbreviations: ADL. = Activities of Daily Living**; AFED** = Age Friendly Emergency Department; **APP - GED Specialist** = Advance Practice Provider, Geriatric Emergency Department; **AWOL+i** = Age, World, Orientation, Illness and Informant Screening Tool**; bCAM** = Brief Confusions Assessment Method Delirium Screen**; CODE DICE** = Overhead page for emergent behaviors of dementia or delirium which leads to huddle of professionals to develop patient centered responses; **EHR** = Electronic health record; **EPIC** = Epic Electronic Health Record; **ESI** = Emergency Severity Index; **GED-SS** = GED Screning Score; **GENIE** = Geriatric Emergency Nurse; **IADL** = Instrumental Activities of Daily Living; **ISAR** = Identification of Seniors At Risk Screening Tool; **Kinder 1** = ED patient fall risk assessment tool; **MAC** = UCSF Memory and Aging Center; **MMSE** = Mini Mental State Exam; **MOCA** = Montreal Cognitive Assessment; **PCT** = Patient Care Technician; **SLUMS** = Saint Louis University Mental Status examination; **STEADI** = Stopping Elderly Accidents, Deaths, and Injuries; **TUG** = Time Up and Go fall risk assessment.

| **GED Intervention Design and Care** | **KPSF** | **UCSF** | **ZSFG** |
| --- | --- | --- | --- |
| Eligibility Criteria for GED Program | 70+ ESI ≥ 2, with GED-Screening Score (GED-SS) flag OR clinical team GED order | 65+ ESI ≥ 2, with ISAR ≥ 2 (through April 2024) OR Frailty questionnaire (gestalt; May 2024 onwards) | 65+ ESI = 2 to 4, with ISAR ≥ 2 |
| GED program trigger/Signal | Electronic health record GED-SS (EHR) flag | AFED Consult order | EHR flag with silver age box for patients 65+ and ESI |
| Who can manually trigger patient for GED program? | Physician, RN, AFED team | Physician, APP, RN, AFED team | Physician, RN, APP, AFED team |
| **GED Care processes** |  |  |  |
| Protocol or care process to standardize and minimize urinary catheter use. | Hospital policy/protocol | Hospital policy/protocol | ED nursing policy/protocol |
| Protocol or care process to minimize NPO status and promote access to appropriate food and drink. | GED Orderset, Patient Care Rounding, GENIE | APP - GED Specialist | ED nursing policy/protocol and physician order |
| Protocol or policy to minimize use of physical restraints and promote use of trained companions or sitters instead. | Hospital policy/protocol | AFED care process | ED nursing policy/protocol |
| **Medication Safety and Orders** |  |  |  |
| Care process for medication reconciliation to be performed by pharmacist or pharmacy technician. | GED Pharmacist | AFED Pharmacist & Pharmacy Technician | Pharmacy technician for admitted patients |
| Guidelines to minimize potentially inappropriate medication use. This could be through an ED-based pharmacist or through a hospital-specific or other list of potentially inappropriate medications (PIMs) or dosing | GED Pharmacist, EHR best practice advisories | AFED Pharmacist & AFED APP | Beers Criteria for bedside nurse + ED pharmacist |
| Guidelines for safe pain control including multi-modal | GED Pharmacist | AFED Pharmacist | Customized Geriatric order set |
| options for mild, moderate, or severe pain. | | | with preselected medications and doses opened by default for all patients $\geq 65$ years old. |
| Development and implementation of at least three order sets for common geriatric ED presentations developed with particular attention to appropriate geriatric medications and dosing and management plans (e.g. delirium, hip fracture, sepsis, stroke, ACS). | GED Pharmacist | AFED Pharmacist | ZSFG Hip fracture protocol; Fall and Physical Therapy consultation guidelines; Vestibular screening guidelines; Pain Protocol; Guidelines for Palliative Care Consultation |
| **ED Specialty Consultation Resources** |  |  |  |
| Care process for accessing palliative care consultation in the ED | Physician consult order | Physician consult order | Physician consult/order |
| Care process for accessing geriatric psychiatry consultation in the ED. | Physician consult order (general psychiatry) | Physician consult/ order (general psychiatry) | Physician consult/order; Psychiatric Emergency Services, behavioral psych attending / consultation MD available 24 hours per day |
| Care process to guide the use of volunteers in the care of older ED patients. | KPSF Volunteer services working with the GENIE | UCSF Volunteer Services | ZSFG ED AFED volunteer handbook for volunteers.<br>AFED Volunteer program |
| **GED Screens and Assessments** |  |  |  |
| Protocol for structured delirium screening with an established tool, with appropriate follow-up actions | bedside RN (bCAM) | APP - GED Specialist (bCAM) | Bedside Nurse using Nursing Delirium Screening |
| based on screening results.<br>Example tools include the DTS followed by the bCAM, 4AT, or others. |  |  | Scale (NU-DESC) (x1/shift) |
| Protocol for structured cognitive impairment screening with an established tool, with appropriate follow-up actions based on screening results. Example tools include the Ottawa 3DY, mini-cog, SIS, short Blessed test, or others. | GENIE (MiniCog, 2/2023-12/2024) | APP - GED Specialist (MiniCog; AWOLi, MOCA, SLUMS, MMSE) | Patient Navigator using MiniCog |
| Protocol for structured assessment of functional screening with an established tool, with appropriate follow-up actions based on screening results. | Automated GED Screening Score Algorithm, GENIE geriatric assessment, TUG test | APP - GED Specialist (ISAR, ADL, IADL) | Bedside nurse performs ISAR; Patient navigator screens for ADL and iADL using Katz and Lawton tools. AFED Social Worker also has access to Katz and Lawton tools. |
| Protocol for structured falls and mobility screening using an established tool, with appropriate follow-up actions based on screening results. Example tools include the fall risk Timed Up and Go (TUG), or other tools. | Technician (TUG) | APP - GED Specialist (STEADI, ADL/IADL) | Kinder1 tool; TUG assessment |
| **Follow Up Care Systems** |  |  |  |
| Health system outpatient follow-up processes | Automatic notification of KP PCPs if patient had ED visit<br>No follow-up scheduled for non-KP members | Outpatient follow-up referrals requested by ED<br>Appointments may be within 90 day but are not guaranteed | Follow-ups referrals only available for patients with government supported insurance (i.e., Medi-care/cal/caid) or self-pay (no commercial insurance referrals permitted for outpatient care) |
|  |  |  | Specialty referrals only through PCP |
| Memory Clinic Referral | PCP notified for follow up MoCA by GENIE; referral to memory clinic if appropriate | Two ED referral processes to MAC: 1. Direct referral with navigation assistance (5/1/22 - 2/28/23); 2. Referral through PCP and then MAC (3/1/23 onward) | PCP notified for follow up reassessment and referral to memory clinic |
| Geriatric Clinic Referral | N/A | Direct referrals from the ED permitted | Direct referrals from the ED permitted |
| Neurology Clinic Referral | Direct ED referral to clinic via neurology consult.<br>Dementia evaluation only for <65 or those with atypical features | Direct ED referral to general Neurology for non-memory/ cognitive issues | Specialty referrals only through PCP.<br>Direct referrals from ED to Neurology permitted |
| PCP referrals | Automatic notification of KP PCPs if patient had ED visit<br>All hospitalized patients scheduled for PCP follow-up at discharge | PCPs listed in the chart, sent notification of ED visit (AFED note will also be sent if applicable)<br>Patients without PCP, AFED program can request general referral to UCSF Primary Care | All ED patients instructed to make PCP follow-up appointments<br>AFED patient navigator assists with scheduling PCP follow-up |
| Social Services Programs | Social Worker consult available in the ER 7 days a week who can place referrals | Community Resources Link and Find Help Link in EPIC (the community | Adult Day Health, Enhanced case management, meal delivery services, |
|  | and linkage to community resources | referrals auto-populate the after-visit summary whether admitted or discharged) | transportation, and home safety checks.<br>Please see paragraph below for in-depth ZSFG AFED Social Services Programs |
| Other: Comfort and Behavioral Management | Sensory Cart, Patient Care rounding, volunteers | Sensory Cart; music; calming kits; shared regular rounding by ED nurses and PCTs; preferential boarding; CODE DICE; volunteer services | Sensory Cart |

### Reach of GED programs for dementia patients

To describe and understand the proportion of ED patients with dementia potentially impacted by the GED programs at each of these EDs, sites defined the “reach” of their programs using criteria once their GED programs were staffed, trained, and at full implementation. **Table 5** provides summary rates of GED programs at each site and the number of dementia patients potentially reached by GED care.

**Table 5:** Intervention reach of GED program ED encounters with dementia diagnoses.

| Intervention | KPSF<br>2023 | UCSF<br>(4/2021-12/2024)(3.5<br>years), | ZSFG<br>(August 1, 2024<br>to July 31, 2025) |
| --- | --- | --- | --- |
| Site Definition of<br>“Reach”/inclusion criterion | • bCAM<br>AND<br>• TUG test<br>AND<br>• GENIE<br>OR<br>Pharmacist note<br>In patients 70+ years in age, ESI ≥ 2, with GED-SS flag or GED flag through manual order. | • AFED pharmacy consult<br>OR<br>• AFED social work consult note<br>OR<br>• AFED mCGA consult note<br>In patients 65+, ESI ≥ 2, with AFED Consult order while in ED. | • Patient Navigator<br>OR<br>• Social Work<br>OR<br>AFED Physical Therapist note<br>In patients 65+ with ED encounters ≥ 3 hours between 9a-4p, M-F, and ISAR ≥ 2. |
| Number of **dementia** encounters “reached” by GED program | 475 | 1704 | 230 |
| Estimated number of **dementia encounters eligible** for GED program during periods available | 2,849 | 4,412 | 1,145 |
| % Of ED encounters w/ dementia/MCI diagnosis “reached” by GED programs | **16.67%**<br>(n=475/2,849) | **38.6%**<br>(n=1,704/4,412) | **20.1%**<br>(n=230/1,145) |

#### Kaiser Permanente San Francisco (KPSF)

Older adults 70+ years of age with an Emergency Severity Index (ESI) score ≥ 2 who were flagged by the GED Screening Score or manually flagged by ED provider or staff were eligible to receive GED care. We considered patients reached by the GED program if they had both a delirium screen (bCAM) and mobility screen (TUG test) recorded during the ED visit and if there was a GeriCNS or Pharmacist consult note documented in their record.

#### University of California San Francisco (UCSF)

Those with cognitive impairment or dementia were targeted for AFED interventions with an AFED consult order (generally patients 65+ with ESI ≥ 2). In general, an AFED consult order occurred when a person was considered frail based on function, cognition, or social connectedness. A consult note from either the AFED pharmacist, social worker, or NP were considered evidence of having been reached by the AFED program. In each case, these notes in the EHR represented not only screening and assessment but also clinical recommendations for care plan improvements.

#### Zuckerberg San Francisco General (ZSFG)

At ZSFG, patients 65 years or older with an ESI between 2 and 4 who screen positive with an ISAR score ≥ 2 are highlighted on the ED tracking board with a silver background in the age box. The patient navigator and social worker review the charts of those patients highlighted on the tracking board and round in the ED, connecting with the providers and nursing staff to prioritize patients for evaluation. The AFED physical therapist also reviews charts and speaks with the ED team to identify patients who would benefit from their expertise. The providers and nursing staff may also order a consult to the AFED team through the EHR (EPIC). The patient navigator screens for cognitive impairment with the MiniCog screening instrument. If the patient has a pre-existing diagnosis of dementia recognized in the emergency department, a Mini-Cog screening is not performed. A consult note from either the AFED patient navigator, social worker, or physical therapist were considered evidence of having been reached by the AFED program.

## RESULTS

Three San Francisco EDs implemented age-friendly GED care programs to improve the quality of emergency care that persons living with or at risk of cognitive impairment or dementia experience during and after an ED visit. Older adults represented more than 30% of ED visits at two hospitals (33% at KPSF, 36% at UCSF, 17% at ZSFG). Populations were diverse in race and ethnicity with White patients ranging from 23% at ZSFG to 50% at UCSF, Black patients ranging from 8% at UCSF to 20% at ZSFG, Asian patients ranging from 26% at ZSFG to 31% at KPSF. Hispanic patients ranged from 8% at UCSF to 25% at ZSFG. Proportion of older ED patients on Medicare Fee-for-Service (FFS) ranged from 4% at KPSF to 81% at ZSFG, yet 86% of KPSF patients were covered by managed care medical insurance.

For each ED, a significant proportion of their ED visit encounters made by patients 65+ years of age also had dementia or cognitive impairment histories (14.7% of ZSFG, 25.4% of KPSF, and 23.2% UCSF ED patients had previously diagnosed dementia or cognitive impairment conditions in their diagnosis histories). Of these patients, 17% at KPSF, 39% at UCSF, and 20% at ZSFG received care by their respective GED programs. Resource and staffing availability of each program likely reduced maximal reach of the programs for all eligible patients, whereby even more patients with dementia would have likely received care.

### Commonalities in GED dementia care programmatic implementation

#### Screening and Assessment

All three EDs use screening and assessment tools to identify patients at risk of cognitive impairment and dementia. All three EDs included delirium and cognitive function assessments. For delirium screening, two used the bCAM, while one used the NuDesc. All utilized the Mini-Cog for cognitive function screening, while one also completed the MoCA, Saint Louis University Mental Status Exam (SLUMS), and Mini-Mental State Examination (MMSE) for cognitive assessment testing.

#### Data Capture

Each program captured assessment data in patients’ medical records, which is crucial for monitoring changes and maintaining continuity of care.

#### Community Engagement

Each ED worked with community-based organizations to facilitate resources for patients and care partners’ post-discharge care, ensuring a smoother transition of care.

### Differences in GED dementia care programmatic implementation

#### Accreditation Status

Programs started in different years and have achieved varying levels of GED accreditation status in recognition of their GED care. UCSF achieved Level 1 GED accreditation in 2021, KPSF achieved Level 1 in 2024, and ZSFG received initial Level 3 accreditation in 2024, with a recent upgrade to Level 2 in 2025.

#### Patient Population and Focus

KPSF focuses on patients aged 70 and above, whereas UCSF and ZSFG target those aged 65 and above.

#### Resource Allocation and Workflow

KPSF employs an automated risk prediction model; UCSF uses embedded NPs to conduct mCGA; ZSFG relies heavily on navigators and social workers.

#### GED Program Reach

The reach and capacity of these GED programs vary significantly, with differing levels of patient engagement and service provision based on available professionals and operational hours.

## DISCUSSION

The models of care implemented by KPSF, UCSF, and ZSFG showcase diverse approaches to enhance emergency care for persons living with dementia or at risk for cognitive impairment. Differences by program show how each developed, adopted, and adapted programs unique to the needs and characteristics of their health system, community resources, and most importantly, their patient populations. Each program adopted best practices aligned with the common goal of improving emergency care for older adults, particularly those with cognitive impairment. KPSF stands out with its use of an automated risk prediction model, enabling early identification of at-risk older patients with a GED Screening Score. UCSF benefits from embedded NPs with specialized training. ZSFG leverages navigators and social workers for comprehensive patient support. These strategies highlight the potential impact of tailored interventions based on institutional resources and patient demographics.

Across the three EDs, several best practices are commonly implemented to enhance emergency care for patients with dementia. Each program integrates screening and assessment tools designed to identify patients at risk of cognitive impairment, ensuring early intervention and tailored care. Additionally, these hospitals emphasize the importance of data capture, whereby assessment findings are recorded in patients’ medical records to facilitate continuity of care and informed decision-making during transitions between healthcare providers.

Community engagement is a shared strength among the three programs, with each working collaboratively with local organizations to ensure seamless transitions and ongoing support for discharged patients. All three sites worked with community-based organizations to provide resources for patient post-discharge care, promoting smoother transitions and ongoing support for older adults within their communities. These collaborations underscore the importance of a holistic approach to dementia care within the emergency setting. Collaborations that extend beyond acute medical care are necessary to ensure continuous care beyond immediate medical interventions.

All three programs align their efforts to develop interdisciplinary approaches, integrating geriatric medicine, emergency medicine, pharmacy, case management, and social work, among other specialties, to foster comprehensive and sustainable care models. Best practices in geriatric medicine and dementia emergency care models are interprofessional, requiring extensive partnerships to support the care coordination not only within the hospital, but into the community. While there were differences in operational specifics, each hospital creatively utilized community partnerships or programs. KPSF utilized the power of their integrated healthcare system data for patients to develop a screening score. UCSF partnered with the Division of Geriatrics to bring NP consults into the ED. ZSFG created a new role of a non-clinician patient navigator trained to deploy geriatric and cognitive-functional assessments.

Despite the best practices, the implementation of these models also presents several challenges. One significant issue is the relatively low number of patients screened, which is in part due to the limited availability of active ED assessment professionals across all programs. This limitation restricts the ability or programs to comprehensively address the needs of all potential dementia ED patients. Furthermore, cognitive impairment and dementia often cannot be diagnosed accurately unless a patient is at baseline, collateral information is available, a decline in functional status can be documented, and a wide differential diagnosis is considered. Nonetheless, the ED setting, despite the challenges of screening and varied professional expertise, may be an important setting for more efficiently recognizing and responding to those with cognitive impairment, especially when integrated with outpatient and community supports. The referral process and supports for post-emergency care is also variable and depending on health system integration of outpatient clinics and permissible care coordination. The ease or complexity of referral processes and care coordination may significantly impact how well older adults or PLWD are able to navigate follow-up care. As GED programs gain traction and demonstrate value, sustainable resources and prioritization of support for such initiatives will advance scalability and broader applicability. While each program has made strides in implementing comprehensive care strategies, there remains a need for more robust data integration systems to streamline processes and improve patient outcomes consistently.

Overall, the KPSF, UCSF, and ZSFG programs illustrate the importance of adopting innovative, age-friendly approaches tailored to the needs of patients living with or at risk for cognitive impairment within the ED. The lessons learned from these programs emphasize the value of integrating specialized care models into ED settings, encouraging continuous quality improvement, and fostering community partnerships to enhance patient care. As these models evolve, addressing existing limitations and exploring avenues for resource optimization will be critical in scaling these initiatives and improving outcomes for cognitively impaired geriatric patients across diverse healthcare systems.

## Data Availability

KP: All data produced in the present study are available upon reasonable request to the authors.
UCSF: All data produced in the present study are available upon reasonable request to the authors.
ZSFG: All data produced in the present study are available upon reasonable request to the authors.

## Author Contributions

NFD, MDD, JH, KAH, EDI, TJ, MT, JS designed the Geriatric ED programs. HA, DM, DFT, UH drafted the manuscript. All authors contributed substantial revisions. We acknowledge Francesca Long for her significant contributions to this work.

## REFERENCES

1 Cairns C, Ashman J, K K. Emergency Department Visit Rates by Selected Characteristics: United States, 2022. NCHS Data Brief 2024;503(August 2024).

2 Hwang U, Morrison RS. The Geriatric Emergency Department. J Am Geriatr Soc 2007;55:1873–1876.

3 Fang M, Hu J, Weiss J, et al. Lifetime risk and projected burden of dementia. Nat Med 2025 (In eng). DOI: 10.1038/s41591-024-03340-9.

4 Kent T, Lesser A, Israni J, Hwang U, Carpenter C, Ko K. 30-Day Emergency Department Revisit Rates among Older Adults with Documented Dementia. J Am Geriatr Soc 2019;doi: 10.1111/jgs.16114.

5 LaMantia MA, Stump TE, Messina FC, Miller DK, Callahan CM. Emergency department use among older adults with dementia. Alzheimers Dementia 2016;30(1):35–40.

6 Gerlach LB, Martindale J, Bynum JPW, Davis MA. Characteristics of Emergency Department Visits Among Older Adults With Dementia. JAMA Neurol 2023;80(9):1002–1004. (In eng). DOI: 10.1001/jamaneurol.2023.2244.

7 Seidenfeld J, Runels T, Goulet JL, et al. Patterns of emergency department visits prior to dementia or cognitive impairment diagnosis: An opportunity for dementia detection? Acad Emerg Med 2023 (In eng). DOI: 10.1111/acem.14832.

8 Seidenfeld J, Dalton A, Vashi AA. Emergency department utilization and presenting chief complaints by Veterans living with dementia. Acad Emerg Med 2023;30(4):331–339. (In eng). DOI: 10.1111/acem.14686.

9 Hwang U, Shah MN, Han JH, Carpenter CR, Siu AL, Adams J. Transforming Emergency Care for Older Adults. Health Aff 2013;32(12):2116–21.

10 CMS Centers for Medicare & Medicaid Services. FY 2025 Hospital Inpatient Prospective Payment System (IPPS) and Long-Term Care Hospital Prospective Payment System (LTCH PPS) Proposed Rule—CMS-1808-P Fact Sheet. (https://www.cms.gov/newsroom/fact-sheets/fy-2025-hospital-inpatient-prospective-payment-system-ipps-and-long-term-care-hospital-prospective2025.)

11 American College of Emergency Physicians. GEDA care processes list. (https://www.acep.org/siteassets/sites/geda/media/documnets/geda-care-processes-list.pdf).

12 Adler-Milstein J, Rosenthal S, Thrombley R, et al. Outcomes Associated With an Age-Friendly Emergency Department Intervention. Ann Emerg Med 2025;in press.

13 Hauser K, Swartzberg J, Schlessinger D, et al. Development of a predictive model for identifying high-risk older adults for geriatric emergency department screening. J Am College Emerg Physician Open in revision.

14 Allan L, Wheatley A, Smith A, et.al. An intervention to improve falls in dementia: the DIFRID mixed-methods feasibility study. Southampton (UK): NIHR Journals Library, 2019.

15 Adler-Milstein J, Gopalan A, Huang J, Toretsky C, Reed M. Patterns of Telemedicine Use in Primary Care for People with Dementia in the Post-pandemic Period. J Gen Intern Med 2024;39(15):2895–2903. (In eng). DOI: 10.1007/s11606-024-08836-1.

16 Healthcare Cost & Utilization Project. Clinical Classifications Software Refined (CCSR) for ICD-10-CM Diagnoses. In: Agency for Healthcare Research and Quality (AHRQ), ed.2024.

17 McCusker J, Bellavance F, Cardin S, Trepanier S, Verdon J, Ardman O. Detection of older people at increased risk of adverse health outcomes after an emergency visit: the ISAR screening tool. J Am Geriatr Soc 1999;47(10):1229–37. (In eng) (http://www.ncbi.nlm.nih.gov/entrez/query.fcgi?cmd=Retrieve&db=PubMed&dopt=Citation&list_uids=10522957).

18 Counsell SR, Callahan CM, Buttar AB, Clark DO, Frank KI. Geriatric Resources for Assessment and Care of Elders (GRACE): a new model of primary care for low-income seniors. J Am Geriatr Soc 2006;54(7):1136–41. (In eng). DOI: 10.1111/j.1532-5415.2006.00791.x.

19 Ramesh T, Kadakia K, Moura L. Transforming Value-Based Dementia Care-Implications for the GUIDE Model. JAMA Intern Med 2024;184(3):237–239. (In eng). DOI: 10.1001/jamainternmed.2023.7669.

20 Degesys NF, James T, Erickson M, Hardy J, Rogers S. A Novel Approach to Addressing Neuropsychiatric Symptoms in the UCSF Emergency Department: Code DICE. Acad Emerg Med. 2026 May;33(5):e70310. doi: 10.1111/acem.70310. PMID: 42132352; PMCID: PMC13174910.

21 Windon C, S P, Erickson M, et al. Navigating Barriers to Dementia Specialty Care Among Vulnerable Populations. Insight from a Multidiscipline Care Navigation Team. J Alzheimers Dis Reports 2024;8:1–6. DOI: DOI: 10.1177/25424823241308456.

